# Deep plasma proteomics identifies and validates an eight-protein biomarker panel that separate benign from malignant tumors in ovarian cancer

**DOI:** 10.1101/2024.10.10.24315232

**Authors:** Mikaela Moskov, Julia Lindberg Hedlund, Svetlana Popova, Maria Lycke, Emma Ivansson, Anna Tolf, Ulf Gyllensten, Karin Sundfeldt, Karin Stålberg, Stefan Enroth

## Abstract

Ovarian cancer has the highest mortality of all gynecological cancers and in symptomatic women, surgery is commonly used as final diagnostic. Available literature indicates that women with benign tumors could often be conservatively managed but accurate molecular tests are needed for triaging where gold-standard imaging techniques are inconclusive or lacking. Here, we analyzed 5416 plasma proteins in two independent cohorts (N=171+233) with symptomatic women that have been surgically diagnosed with benign or malignant tumors. Using one cohort as discovery, we compared protein levels of benign tumors with early stage (I-II), late stage (III-IV) or any stage (I-IV) ovarian cancer. In this analysis, 327 associations, corresponding to 191 unique proteins, were identified out of which 326 (99.7%) replicated. The 191 proteins were compared with their corresponding tumor gene expression in the replication cohort and only 11% (21/191) were found to have significant correlation. Protein-protein correlation networks were generated and 62 of the 191 proteins were highly correlated with at least one other protein, suggesting that many of the observed associations could be secondary effects. Multivariate models were trained using the discovery cohort including a fixed cut-off for malignancy. In the replication cohort, an eight-protein model achieved an AUC of 0.96 corresponding to 97% sensitivity at 68% specificity. For early-stage tumors, the sensitivity was estimated at 91% at 68% specificity compared to 85% and 54% for CA-125 alone. Our results indicate that up to one third of benign cases could be identified by molecular measures thereby reducing the need for diagnostic surgery.

**One Sentence Summary:** Plasma proteomics for separation of benign and malignant tumors in ovarian cancer.

## INTRODUCTION

Ovarian cancer is the deadliest of all gynecological cancers and the 8^th^ most common female cancer overall with over 300 000 new cases and 200 000 deaths per year*(1)*. Discovery is mainly symptom driven or by incidental finding*(2)*, and there are no biomarkers available today that could justify general screening*(3)*. The common late-stage diagnosis leads to an overall 5-year survival of ovarian cancer of only 30-50%. However, if the cancer is detected in stage I, close to 90% of patients can be cured, while patients with spread cancers detected in stage III or IV has a 5-year survival rate of less than 30%*(4)*. Few molecular biomarkers are clinically used today to complement imaging examinations, but none have sufficient accuracy to be used for screening nor for accurate diagnostics in symptomatic women. Recent investigations based on single cell mRNA sequencing have shown patient-specific gene expression patterns, specific changes in both gene-expression in the surrounding tumor micro-environment, as well as cell-type composition in relation to tumor progression*(5)*. The plasma proteome could potentially be a reflection of such protein expression changes in any of the affected cell types. Differentially expressed circulating proteins biomarkers as detected in plasma could therefore stem from the actual tumor cells as well from a larger variety of surrounding cell types directly or indirectly affected by the progressing tumor*(6)*.

When diagnostic imaging indicates adnexal mass, surgery is often necessary for a final diagnose. Most of the adnexal mass cases are of benign nature*(7, 8)* and there are indications that these could be conservatively managed, e.g. without surgical intervention, or by choosing less invasive surgical procedures with low risk of complications*(2)*. Surgical intervention, by itself, is not risk-free and surgery related complications have been reported in 3.5% to as high as 15% of the women with benign adnexal mass*(2, 9)*. Imaging techniques can achieve high accuracy in separating benign from malignant conditions*(10)*, but as noted in a recent Cochrane review*(11)*, the bulk of the present literature evaluating the imaging techniques is based on studies conducted in tertiary settings, and the clinical setting has both a significant impact on the performance and the cost-benefit for the health care system. In addition to imaging techniques which commonly require highly trained personnel to achieve the highest accuracy, molecular preoperative tools that accurately separate benign from malignant cases could help in reducing referrals to tertiary centers and unnecessary surgical interventions, thus minimizing potential complications and side-effects such infertility or premature menopause*(9)*. MUCIN-16 (CA-125) is currently the best single molecular biomarker used for ovarian cancer diagnosis in post-menopausal women and in treatment management*(12)*. MUCIN-16, however, has low sensitivity for early-stage cancer and can also be elevated in many benign gynecological conditions in younger women, such as infections, pregnancies, and endometriosis*(12, 13)*, resulting in a high proportion of false positives when discriminating between benign and malignant ovarian cancer tumors. MUCIN-16 levels have also been found to be above the clinically indicative cut-off for ovarian cancer (35 U/mL) in close to half of women with acute pancreatitis*(14)* and in 1 of 20 elderly women with heart failures*(15)*. Combining MUCIN-16 with other biomarkers, including WAP Four-Disulfide Core Domain 2 (WFDC2 or HE4), as in the ROMA Score (Ovarian Malignancy Risk Algorithm) improves the performance. The ROMA score is calculated differently in pre- and post-menopausal women and have been reported with an overall sensitivity of 87.0% at a specificity of 80.9% in pre-menopausal and with 91.1% and 77.2 % in post-menopausal women respectively*(8)*. In early stages, the sensitivities in pre-/post-menopausal women have been reported at 77.8/81.4% at specificities of 80.9/77.2%*(8)*. The lower sensitivity at the indicated cut-off for detection of early-stage ovarian cancer (stages I and II prohibits accurate discrimination of benign and malignant conditions in symptomatic women with the risk of a large proportion of the cancer cases remaining undetected. Several studies have indicated that combining several protein biomarkers into a single test can increase test performance. The OVA1-test, for instance, combines five proteins (Apolipoprotein A1, Beta 2 microglobulin, MUCIN-16, Transferrin and Prealbumin/Transthyretin) and classifies women into categories of high, intermediate or low risk of ovarian cancer. In a multi-center study*(16)* a higher proportion of the individuals predicted to be low risk, e.g. having benign tumors, according to the OVA1-test remained benign during a 12-month follow-up period as compared to using MUCIN-16 alone. A second generation of the OVA1-test, OVERA*(17)*, also combining five proteins (MUCIN16, Transferrin, Apolipoprotein A1, Follicle-stimulating hormone and WFDC2) increased the sensitivity to an estimated 69% at a specificity of 91%. OVERA is reported to have a sensitivity of 88.6% in detecting early-stage cancers (stage I and II) *(17)*. We have previously developed*(18)* and validated*(19)* an 11-protein biomarker panel that outperformed MUCIN-16 in separating benign from malignant conditions at time of diagnose. This panel was constructed based on analyses of up to 983 plasma proteins and achieved sensitivities of 83-88% at specificities of 88-92% at a pre-defined cut-off across two independent validation cohorts with both pre- and post-menopausal women *(18)*. Additional studies of up to 1536 plasma proteins*(20, 21)* and up to 3072 proteins*(22)* have identified multiple additional biomarker candidates for ovarian cancer with promising results, but additional validation is needed before clinic use. These studies indicate that combinations of biomarkers, even without inclusion of MUCIN-16, can achieve high precision in separating benign from malignant conditions, and that large-scale characterizations of the plasma proteome in combination with machine learning represents a promising route for development of novel tests for separation of benign and malignant ovarian tumors. In this study, we aimed to identify and validate single and multiplex biomarkers for separation of malignant and benign ovarian cancer tumors in symptomatic women. To this end, over 5400 plasma proteins in each sample were characterized using high-throughput affinity-based proteomics and RNA-sequencing in corresponding tumor tissue was used to assess tumor gene-activity in relation to the plasma proteins. Finally, we employed machine learning to identify a small biomarker signature consisting of eight proteins that predicts malignancy, and the performance of this signature was validated in a separate cohort.

## RESULTS

### Deep plasma proteome characterization

The plasma proteome of 404 women surgically diagnosed with benign or malignant conditions after suspicion of ovarian cancer was studied using the proximity extension assay (PEA) implemented in the Olink Explore HT assay*(23)*. The samples were collected from two independent Swedish cohorts (Table 1) at time of diagnose, before initiation of treatment. The samples were collected at two different geographical locations, with the samples collected in Göteborg used as discovery cohort and the samples collected in Uppsala as replication cohort. A total of 5416 unique proteins was characterized in each sample. After basic quality control (see Methods) requiring at least 95% detection rates in both individuals and proteins, 5414 proteins in all 404 individuals were included in further analyses. Both cohorts have clinically measured CA-125 (Table 1), and a significant (p < 1.3 x 10^-44^) correlation was found between clinical CA-125 and the MUCIN-16 assay on the Olink Explore HT, with an estimated correlation coefficient of 0.74 (Spearman’s Rho). The protein content on the assay reflects a broad spectrum of biological processes and functions, not only with previous relation to cancer, and in line with this, using all 5414 proteins we observed no clear distinction between benign and malignant diagnosis using the first two PCA dimensions (Figure 1A) in the discovery cohort with a total of 15% of the total variance explained by the first two principal components. A similar pattern was seen when projecting the replication cohort onto the same plane (Figure 1B).

**Fig 1.**
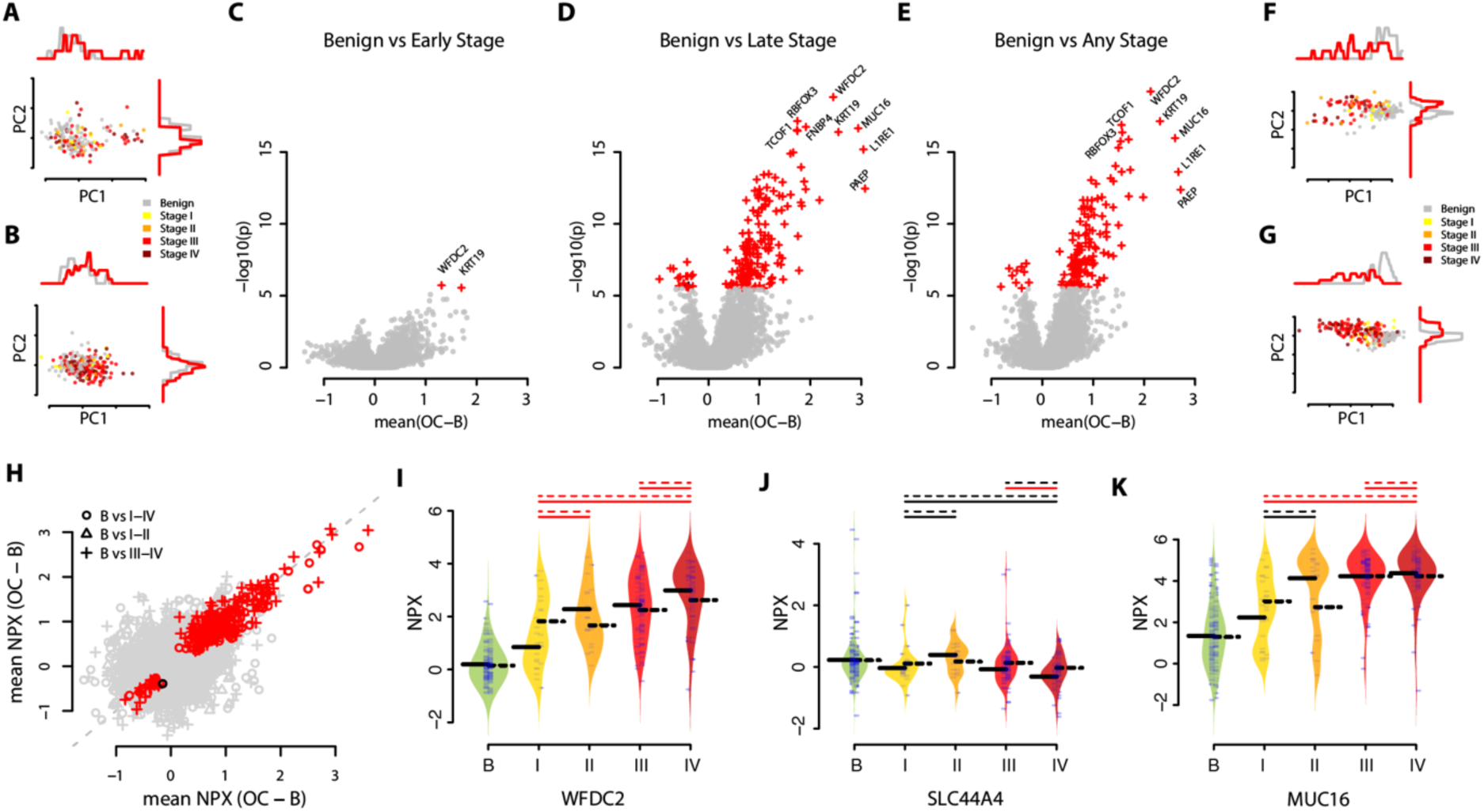
Protein biomarker candidates for ovarian cancer. **(A)** PCA-projection of the discovery cohort using all proteins. Individual samples are labelled based on diagnosis. **(B)** Projection (PCA) of the replication cohort using the same transformation in (A). **(C)** Volcano-plot showing mean difference between early-stage ovarian cancer and benign diagnoses on x-axis and statistical significance (two-sided Wilcoxon ranked sum test) on the y-axis in the discovery cohort. Proteins with significant difference (q-value < 0.05, adjusted with Holm’s method) in the discovery cohort are drawn as crosses and colored by statistical significance in the replication cohort, with red indicating replicated and black not replicated. **(D)** as (C) but for benign diagnoses compared to late-stage ovarian cancer. **(E)** as (C) but for benign diagnoses compared to all stages of ovarian cancer. **(F)** as A) but using significant biomarkers (two-sided Wilcoxon ranked sum test, q-value < 0.05, adjusted with Holm’s method) only. **(G)** Projection (PCA) of the replication cohort using the same transformation in (F). **(H)** Comparisons of the fold changes between cases and controls in the discovery cohort (y-axis) and the replication cohort (x-axis). Proteins with significant difference (q-value < 0.05, adjusted with Holm’s method) are draw as crosses and colored by statistical significance also in the replication cohort with red indicating replicated and black not replicated. **(I)** WCFD2 protein abundance in plasma in relation to diagnose and stage (x-axis). Each ‘beanplot’ show distribution of discovery cohort to the left and the replication cohort to the right. Thick black horizontal lines indicate mean in each group with solid lines for the discovery and dashed lines for the replication cohorts respectively. Thin lines above the beanplots indicate statistical significance between the spanned stages and the benign group with red indicating q-value < 0.05 and black > 0.05. In each set of lines, the dashed line is for the replication cohort and the solid line is for the discovery cohort. **(J)** as (I) but for SLC44A4. **(K)** as (I) but for MUCIN-16 (labelled ‘MUC16’).

**Table 1.** Cohort characteristics.

|  | Cohort | All | Benign | Ovarian Cancer |  |  |  |
| --- | --- | --- | --- | --- | --- | --- | --- |
|  |  |  |  | I | II | III | IV |
| Nr. Samples | Discovery | 171 | 86 | 13 | 11 | 38 | 23 |
|  | Replication | 233 | 81 | 13 | 10 | 80 | 49 |
| Age at Diag.<sup>a</sup> | Discovery | 60.3 (14.6) | 58.1 (16.6) | 62.2 (11.5) | 67.7 (8.7) | 60.3 (12.4) | 63.8 (12.4) |
|  | Replication | 61.5 (12.0) | 58.2 (14.0) | 61.7 (10.5) | 67.6 (8.7) | 62.0 (10.6) | 64.8 (10.3) |
| Age diff pval<sup>b</sup> |  | 0.71 | 0.79 | 0.96 | 0.97 | 0.76 | 0.74 |
| Clin. CA-125<sup>c</sup> | Discovery | 820.4 (1989.4) | 85.8 (181.8) | 107.3 (156.7) | 546.3 (642.4) | 1957.0 (3017) | 2214.8 (2874.3) |
|  | Replication | 1175.3 (2356) | 93.1 (138.4) | 587.7 (862.9) | 1006.8 (1122.6) | 1354.0 (2610.3) | 2309.9 (3176.2) |
| CA-125 diff pval<sup>b</sup> |  | 0.71 | 0.79 | 0.96 | 0.97 | 0.76 | 0.74 |

<sup>a</sup> Reported as mean (std dev) age in the group. <sup>b</sup> Two-sided Wilcoxon ranked test comparing the Discovery and Replication cohorts <sup>c</sup> Clinically measured CA-125 at time of diagnosis, reported as mean (std dev) U/ml in the group.

### Replicated single protein biomarkers candidates for ovarian cancer

Using the discovery cohort, each of the 5414 proteins was compared in three different setups: benign vs early-stage ovarian cancer (stage I and II), benign vs late-stage ovarian cancer (stage III and IV), and benign vs any stage ovarian cancer (stage I, II, III, or IV). Across all 16242 comparisons (5414 proteins x 3 comparisons), we found 327 significant associations (q-value < 0.05, adjusted for multiple hypothesis testing with the Holm’s method) in the discovery cohort (Supplementary Table 1). The 327 associations corresponded to a total of 191 unique proteins with two proteins (Keratin-19 (KRT19) and WAP four-disulphide core domain protein 2 (WFDC2) found to be significantly different in all three comparisons (Figure 1CDE). Each of the 327 associations was then examined using the replication cohort, and 326 (99.7%, Figure 1CDE) remained significant after adjustment for multiple hypothesis testing (Holm’s method). The one protein that did not replicate, Solute Carrier Family 44 Member 4 (SLC44A4), showed a significant association only between late-stage cancer and benign tumors in the discovery cohort (Supplementary Table 1). Using all the 191 proteins that showed a significant difference in the discovery cohort, we observed a trend towards a distinction between benign and malignant diagnoses in the first two PCA dimensions in the discovery cohort with a total of 53% of the total variance explained by the first two principal components (Figure 1F), as well in the replication cohort (Figure 1G) when projected onto the same plane. We next investigated the fold-change between the benign and malignant groups for the 327 associations and found a high similarity (Figure 1H, Pearson’s Rho 0.93, p < 3.0 x 10^-140^), both for biomarkers with a higher expression in those with malignant versus those with benign diagnoses and vice versa (Figure 1H). We also observed different patterns among the protein biomarkers in relation to the cancer stage, with e.g. increasing levels with stage or non-linear plateauing. Figure 1I-K shows the observed distribution of protein levels in benign and malignant diagnoses for both the discovery and replication cohort for three proteins. Figure 1I shows the top ranking hit overall, WFDC2, with increasing levels for stages I-IV compared to benign. Figure 1J shows the only protein that did not replicate, SLC44A4 and Figure 1K shows the patterns observed in MUCIN-16. Note that MUCIN-16 was not significantly different between benign and early cancer stages in the discovery cohort nor the replication cohort. All results for the three comparisons are reported in Supplementary Table 1. In conclusion, we found a large number of potential plasma protein biomarkers out of which over 99% validated in a replication cohort. Not all biomarkers were however showing difference across both early and late cancer stages as compared to benign tumors and few displayed a clear linear relationship with cancer stage.

### Plasma protein biomarkers are in general not correlated with tumor gene expression

The tumor microenvironment is a complex environment and circulating proteins biomarkers could stem from the actual tumor cells or from a larger variety of surrounding cell types directly or indirectly affected by the progressing tumor*(6)*. To assess the possible origin of the plasma proteins here we characterized the gene-expression in 81 tumor samples from the replication cohort with total RNA-sequencing (see Methods). This included samples of both benign (N=10) and malignant tumors (N=71, N stage I-IV: 5, 5, 37, 24). Using the ‘transcript per million reads’ (TPM) score we then calculated the correlation coefficient between tumor mRNA expression and the plasma protein levels. We found non-zero gene expression levels in at least one sample for 5364 of the 5414 proteins (99.1%). When comparing the corresponding tumor gene expression with the plasma protein levels, 33 protein-gene pairs, corresponding to 0.62% (33/5364, Table 2), were found to have a significant correlation in a paired analysis (Spearman’s Rho, multiple hypothesis adjusted q-value < 0.05, Supplementary Table 2). When examining the 191 proteins whose levels were found to be significantly different between benign and malignant tumors, 21 proteins (Table 2), corresponding to 11.0% (21/191), showed a significant correlation with the corresponding tumor mRNA expression. This represents a significantly higher proportion than in the comparison of all proteins examined (p < 6.8 x 10^-20^, binominal test). Across the 191 proteins, both proteins with an increase or a decrease in plasma protein concentration in malignant tumors as compared to benign were found to have both positive or negative correlations with their corresponding tumor gene-expression (Figure 2A). Among the 33 proteins showing a significant correlation between plasma and RNA levels, however, only positive correlations were observed (Figure 2B). When requiring both significant difference in plasma proteins expression between malignant and benign diagnoses and significant correlation with tumors mRNA expression, only positively correlated pairs with higher plasma protein concentrations in malignant compared to benign diagnoses was observed (Figure 2C). Among the top-ranked plasma protein biomarkers in the univariate analysis we found examples of both correlated and non-correlated patterns. For instance, WFDC2 plasma levels were found to be significantly correlated (p < 1.7 x 10^-7^) with corresponding mRNA expression in the tumor (Figure 2DE). On the other hand, the plasma level of The RNA Binding Fox-1 Homolog 3 (RBFOX3) was not correlated (p = 0.32) with tumor mRNA expression (Figure 2FG). Alkaline phosphatase, placental type (ALPP) showed the strongest correlation between protein level and tumor mRNA expression (Spearman’s Rho = 0.70, p-value < 6.0 x 10^-13^, Figure 2H). ALPP did not, however, show any difference in protein expression between malignant and benign diagnoses (Figure 2I). These results indicate that correlation between tumor gene expression and plasma protein level is neither a necessity nor an assurance for a strong univariate plasma protein biomarker for ovarian cancer.

**Fig. 2.**
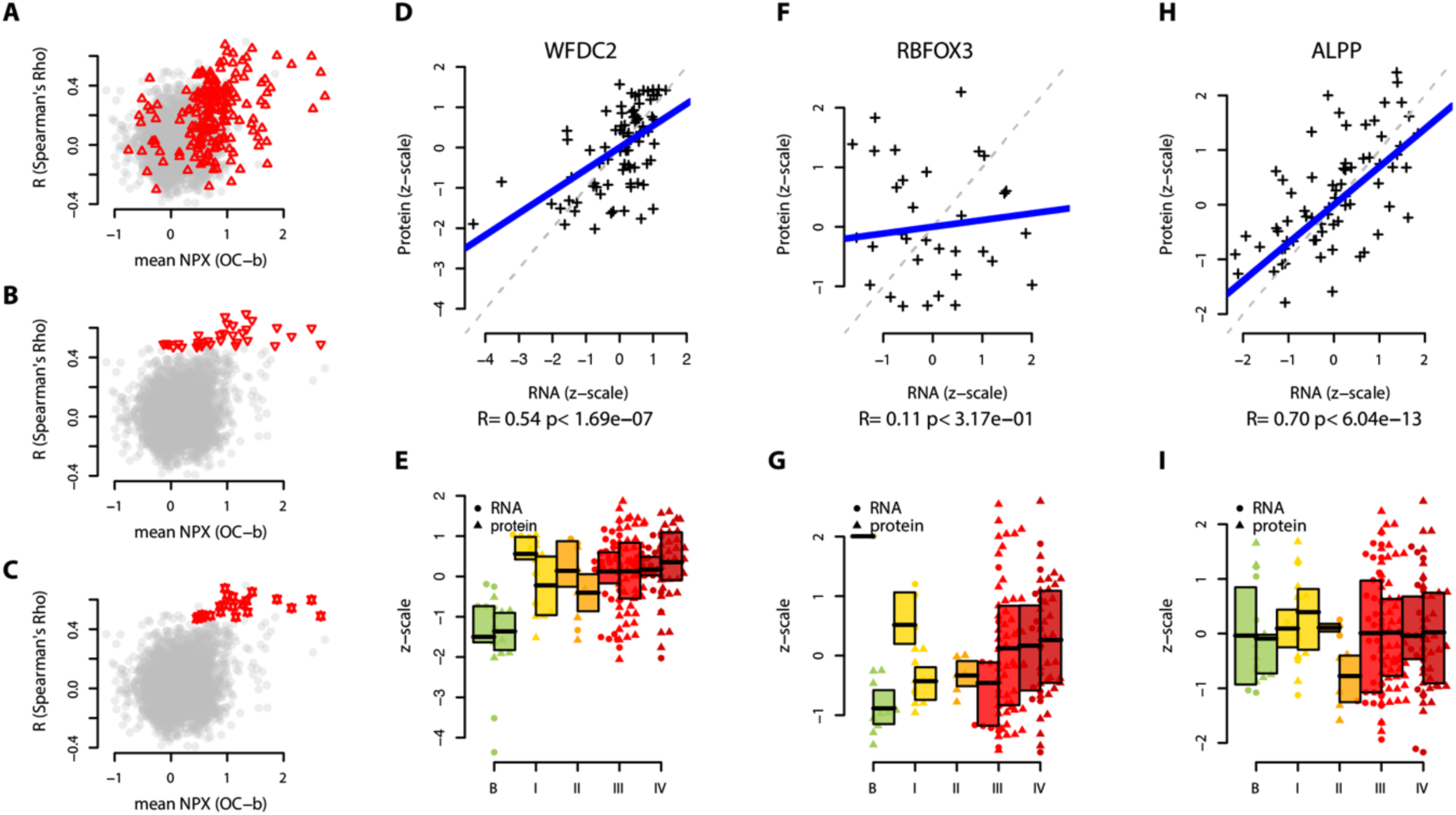
Plasma protein abundance and tissue gene expression. **(A)** Correlation (y-axis, Spearman’s Rho) between plasma protein abundance (NPX) and tissue gene expression (TPM) compared to mean difference between benign and malignant diagnoses (x-axis) for all proteins. Red triangles indicate the significant (two-sided Wilcoxon ranked sum test, q-value < 0.05, adjusted with Holm’s method) difference in the discovery cohort between benign and malignant diagnoses. **(B)** As (A) but red triangles indicate significant correlation (two-sided Spearman test, q-value < 0.05, adjusted with Holm’s method) between plasma proteins and tissue gene expression. **(C)** as A) but red double triangles are overlapping features from (A) and (B). **(D)** Protein plasma abundance levels (y-axis) and tissue RNA expression levels (log2, x-axis) for WFDC2. Both axes are z-scaled to have mean = 0 and unit variance. Labels below the x-axis is the Rho estimate and the corresponding p-value (two-sided Spearman test). The thick blue line represents the Spearman’s correlation coefficient. **(E)** Boxplots with individual protein (right) and RNA abundance (left) levels for benign (labelled ‘B’) and malignant (stages I, II, III and IV) diagnoses for WFDC2 The top and the bottom of the box represent the 25th and 75th percentile and the band inside the box the median value. **(F)** and **(G)** as (D) and (E) but for RBFOX3. **(H)** and **(I)** as (D) and (E) but for ALPP.

**Table 2.**
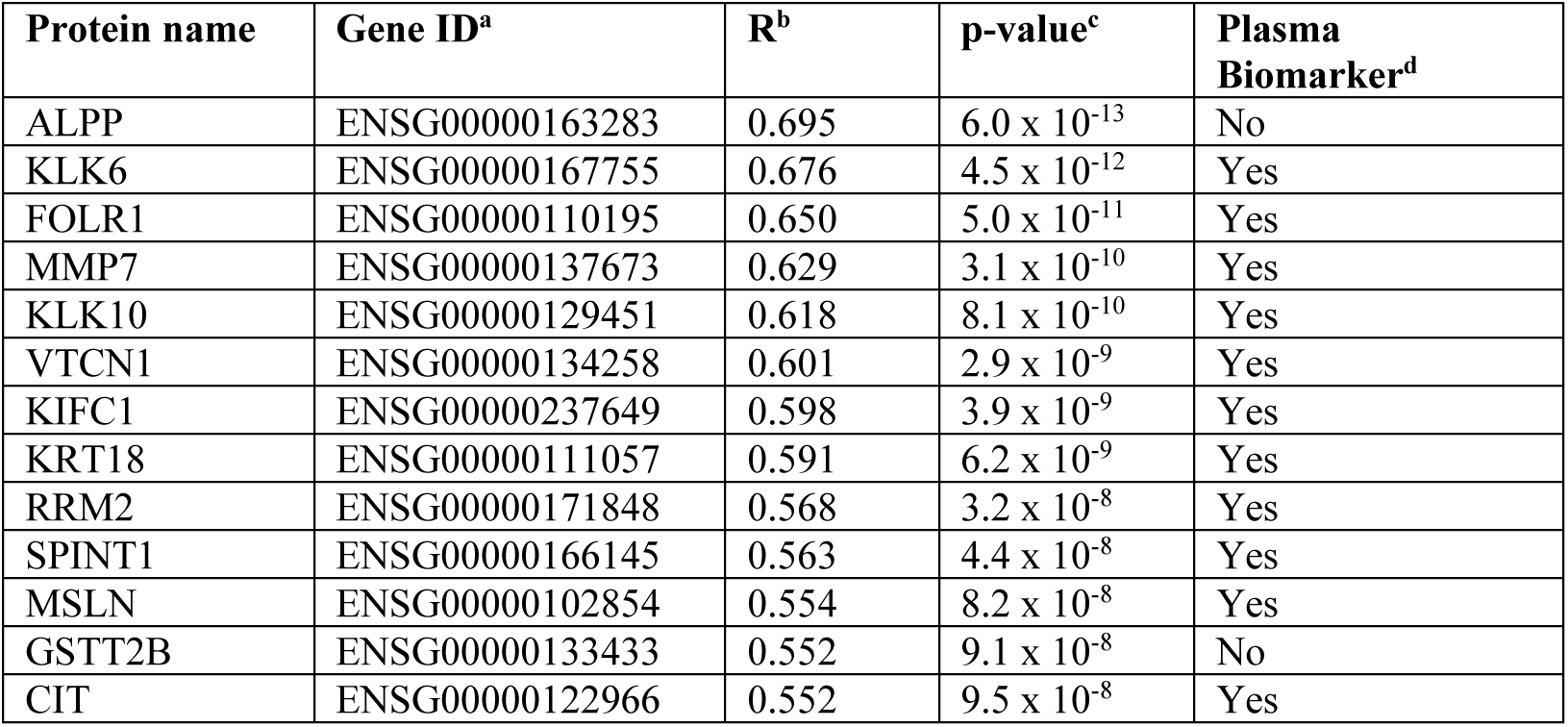
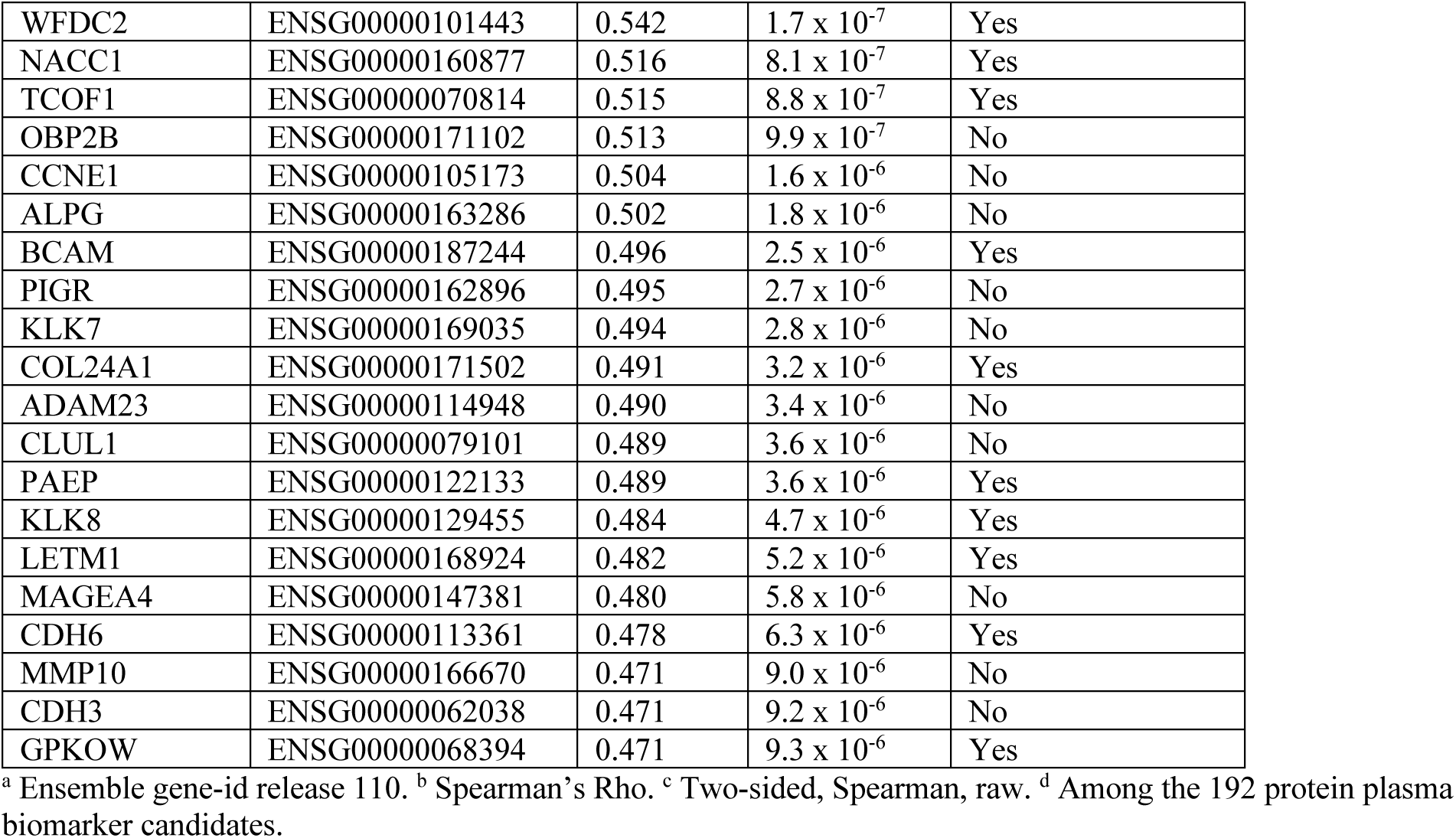
Plasma proteins with significantly correlated tumor RNA expression.

### Correlation of plasma proteome reveals clusters of co-expressed networks

We next investigated protein-protein correlations in relation to ovarian cancer. Starting from the 191 proteins, we calculated protein-protein correlations in relation to all 5414 measured proteins using both the malignant and benign cases of the discovery cohort. We found a total of 106101 significant protein-protein correlations (q-value < 0.05, adjusted for multiple hypothesis testing with Holm’s method) in the discovery data out of which 95.8% (101671) were found to be nominally significant (p-value < 0.05) also in the replication data (Supplementary Table 3). After adjustment for multiple hypothesis testing also in the replication cohort (Holm’s method), 70.9% (75259) remained significant (q-value < 0.05). Based on the significant correlations with an estimated coefficient (Spearmans’ Rho) greater than 0.8, or smaller than -0.8, in the discovery cohort, we then built correlation networks. The highest estimated correlation factor found for MUCIN-16 was 0.72 (with Carboxypeptidase A4 (CPA4), Supplementary Table 3) and MUCIN-16 was therefore not included in the network analysis. The network analysis resulted in 177 individual proteins connected by 315 strong correlations, out of which 62 proteins were among the 191 univariate significant biomarker candidates. In the network analysis, only two of the 315 correlations were not significant also in the replication cohort (Frataxin (FXN) to Proteasome activator complex subunit 2 (PSME2) and Cilia And Flagella Associated Protein 36 (CFAP36) to PSME2). From these general networks, clusters of correlated sub-networks were identified (Methods). This analysis resulted in 15 sub-clusters of interconnected proteins (Figure 3A, Supplementary Table 4). Five of these 15 clusters contained more than 3 proteins, ranging from 17 to 59 proteins each. One of the clusters is shown in detail in Figure 3B, incorporating 29 proteins out of which all but two (Thimet Oligopeptidase 1, THOP1 and Small Nuclear Ribonucleoprotein Polypeptide B2, SNRPB2) were among the 191 proteins found to be significantly different between benign and malignant tumors (Figure 3B). This network also contained 6 proteins (WFDC2, Treacle Ribosome Biogenesis Factor 1 (TCOF1), Folate Receptor Alpha (FOLR1), Leucine Zipper And EF-Hand Containing Transmembrane Protein 1 (LETM1), G-Patch Domain And KOW Motifs (GPKOW) and Keratin 18 (KRT18)) that showed a significant correlation between plasma protein levels and mRNA tumor expression (Figure 3B, Table 2). These results show that, although individual proteins have a significant difference in expression between the malignant and benign groups, several proteins pairs amongst the univariate significant biomarkers are observed to be closely co-expressed. This in turn suggests that the difference between malignant and benign diagnoses could be explained by a subset of the candidates.

**Fig. 3.**
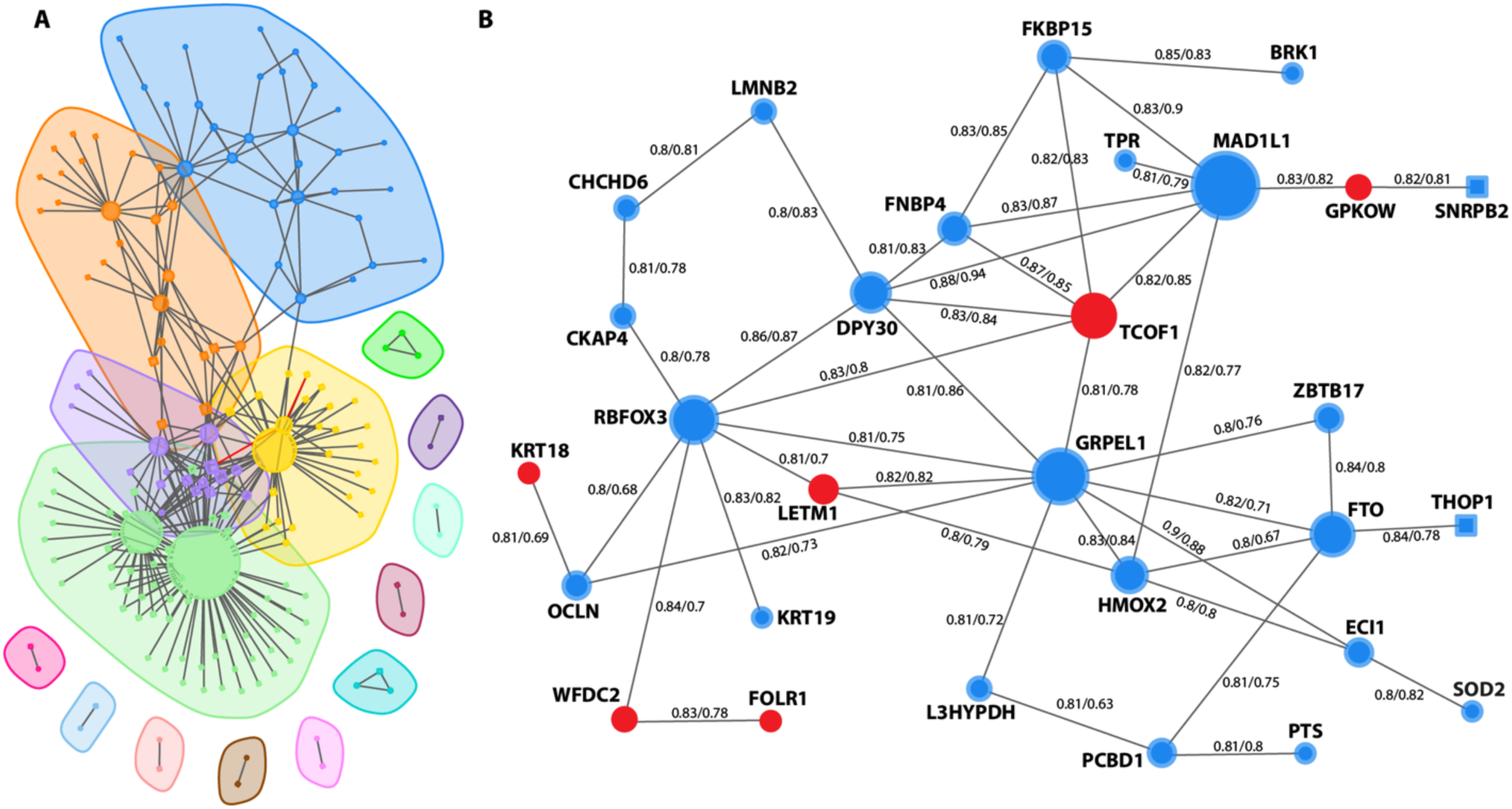
Correlation networks of plasma proteins. **(A)** Clusters of correlated protein-protein pairs as identified in the discovery cohort. Each node represents a unique protein with edges representing correlated protein pairs. Circular nodes correspond to proteins with significant difference between benign and malignant diagnoses in the discovery cohort (two-sided Wilcoxon ranked sum test, q-value < 0.05, adjusted with Holm’s method) while nodes drawn as squares represent non-significant differences. Red edges represent correlations significant only in the discovery cohort. Shaded areas represent generated clusters based on the Leiden algorithm (Methods). **(B)** Detailed representation of one identified cluster from (A). Protein identifiers are written out next to each node and numbers on the edges represent correlation coefficients (Spearman’s Rho) in the discovery cohort and the replication cohort. As in (A), node shape corresponds to statistical significance in case control comparisons. Nodes are colored by RNA-protein correlation where red nodes represent plasma proteins with significant correlation to the corresponding tumor gene expression.

### High-accuracy multivariate risk-score separates benign from malignant tumors

Following our analysis strategy above for the univariate comparisons we built three different multivariate models aiming at separating benign tumors from early stage (I and II), late stage (III and IV) and any stage (stage I-IV) ovarian cancer, respectively. As above, the Göteborg cohort was used as discovery cohort for model development (training) and for selection of a cut-off for predicting malignant condition. The models generate a risk-score on the scale of 0 to 1 and the cut-offs were chosen at a sensitivity of at least 95%. The models were created starting from the 191 proteins with univariate significance in the discovery cohort. As many of the proteins are correlated between themselves, we first used a supervised feature selection (see Methods) to limit the number of proteins in each model. The selected proteins were then used to build predictive models employing distance weighted discrimination*(24)*, a non-linear regression modelling framework for high-dimensional, low sample-size settings. The three models were trained and tuned using the discovery cohort (see Methods). The final model for separating benign vs early stage cancer was based on five proteins (Keratin 19 (KRT19), FOLR1, WFDC2, BRICK1 Subunit Of SCAR/WAVE Actin Nucleating Complex (BRK1) and V-Set Domain Containing T Cell Activation Inhibitor 1 (VTCN1)), the final model for separating benign vs late stage cancer included seven proteins (WFDC2, MUC16, KRT19, TCOF1, Crumbs Cell Polarity Complex Component 2 (CRB2), RBFOX3 and LINE1 retrotransposable element 1 (L1RE1)), and the final model for separating benign vs any stage cancer included eight proteins (WFDC2, KRT19, MUC16, RNA polymerase-associated protein LEO1 (LEO1), TCOF1, Cysteine Rich Secretory Protein 3 (CRISP3), FOLR1 and RBFOX3). In total, twelve proteins were included in any of the models, with only WFDC2 and KRT19 common to all three models (Table 3). We also calculated the relative importance of each protein in the separate multivariate models and found slight differences between the models although similar scores were obtained (Table 3). After training, the models were then evaluated in the replication cohort (Figure 4ABC, Table 4). We found no statistical difference between the performances in the discovery and replication cohort for AUC (all p-values > 0.21, Table 4) nor for the estimated sensitivities (all p-values 0.39, Table 3) or specificities (all p-values > 0.25) at the respective cut-offs as indicated above. We also compared the performance of our models to that of clinically measured CA-125 in the replication cohort (Figure 4ABC). We could observe higher AUCs for all the multivariate models as compared to CA-125, but we found no statistical difference between the respective models and CA-125 (Figure 4ABC, all p-values > 0.14). The model separating any stage from benign conditions is the most applicable in clinical setting, before surgical diagnose and known tumor stage. Therefore, we evaluated the performance of that model when applied specifically to different subgroups in the replication cohort. First, we compared different groups with respect to histology and tumor stage (Figure 4DE). We found no statistical difference in the AUCs for the different histology subgroups as compared to the general performance (all p-values > 0.14), with AUC ranging from 0.90 to 0.99 (Figure 4D). A similar pattern was observed for the subgrouping of tumor stages compared to the general performance (all p-values > 0.24), with AUC ranging from 0.89 to 0.98 (Figure 4E). Specifically, when applying the any stage model to early and late-stage samples separately in the replication cohort, sensitivities of 0.91 and 0.98, respectively, were observed (Table 4). Next, we compared the sensitivities and specificities obtained at the indicated cut-off in our model with the sensitivities and specificities obtained with CA-125 at the commonly used cut-off of 35 U/ml (Figure 4F). The comparison revealed similar point estimates of sensitivities across all samples (model +1.1%) and for the late-stage group (model -0.51%) with a slightly higher point estimate for the early-stage group (model + 6.7%). Looking at the benign cases only, the point estimate of the specificity for the model (67.9%) was 14.1% higher than for CA-125 (53.8%). Using McNemar’s test for non-inferiority in paired samples we found no differences for the sensitivities (p = 1.0) nor for the specificities (p = 0.45) when comparing the predictions made by our model to CA-125. Finally, we analyzed the individual classifications of the any stage from benign model in the replication cohort. At the developed cut-off, the model correctly classified 67.9 % (55 of 81) of the benign samples while miss-classifying 3.3% (5 of 152) of the malignant tumors as benign (Figure 4G). Among these five false negative samples, one sample each was from patients with a stage I, II and III tumor respectively, and two samples were from patients with a stage IV tumor. Four of the five (80%) false negatives were classified as HGSC which is not statistically different (p-value = 1, binomial test) from the proportion of HGSC in the whole replication cohort. The small number of false negatives prevented any additional stratified analysis of the sample group. In summary, our multivariate model displays robust performance in the replication across both tumor subtypes and stages with and estimated 14% higher specificity as compared to CA-125 alone.

**Fig. 4.**
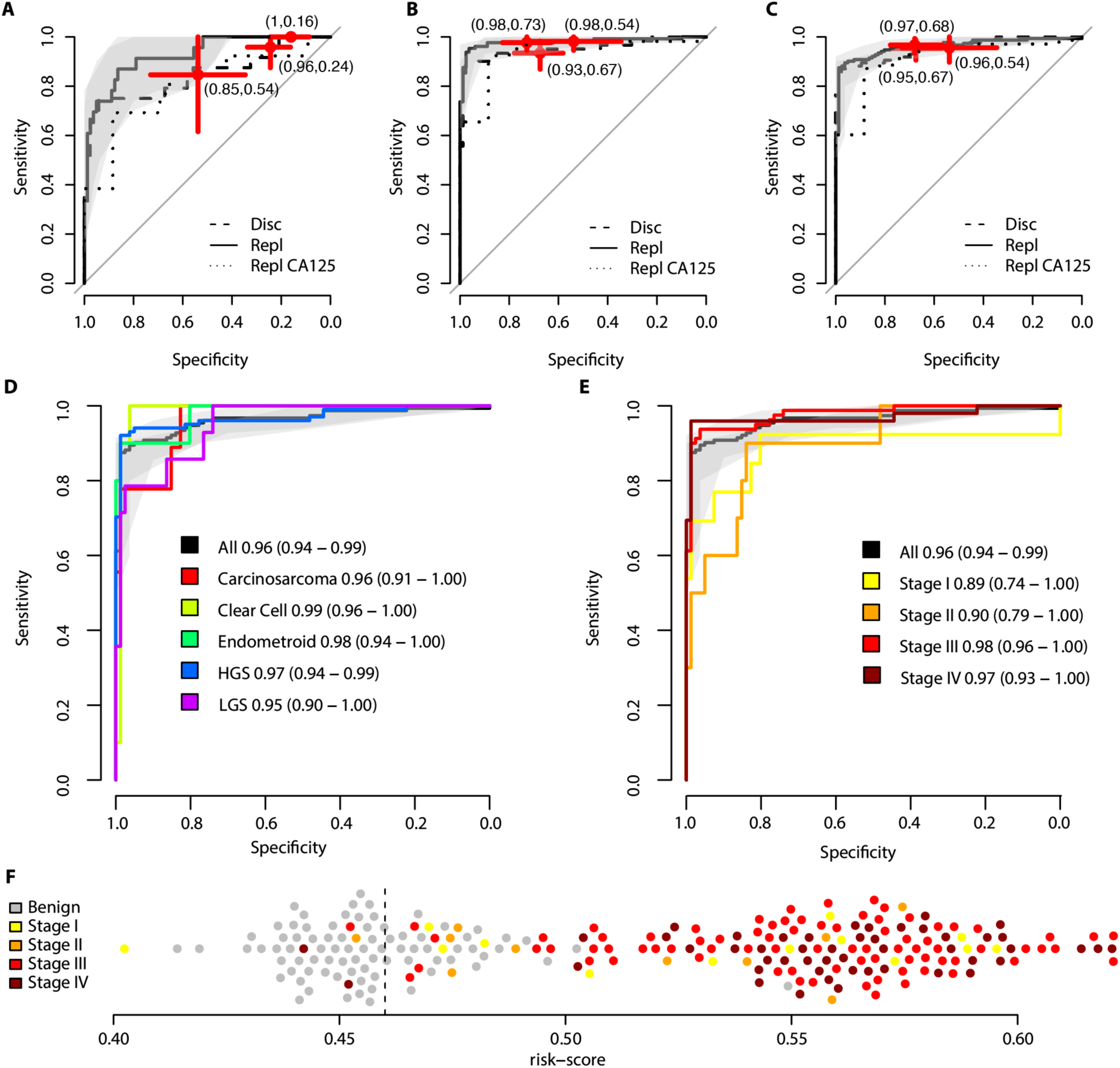
Multivariate prediction models. **(A)** Performance of the benign vs early stage (stages I and II) tumors prediction model. Receiver operating characteristic (ROC) curves are shown for the prediction model in the discovery cohort (dashed line) and in the replication cohort (solid line). The shaded area (grey) corresponds to the 95% confidence interval of the ROC curve in the replication cohort. The red crosses are centered on the point-estimate of the sensitivity and specificity obtained at the cut-off with the horizontal and vertical lines corresponding to the 95% confidence interval of the estimate. The panel also show the performance of clinically measured CA-125 (dotted line) in the replication cohort with the red cross as for the discovery and replication cohort but at a cut-off of 35 U/ml. **(B)** As (A) but for the benign vs late stage (stages III and IV) tumors. **(C)** As (A) but for the benign vs any stage (stages I-IV) tumors. **(D)** ROC curves for stratified analysis of different histology in the replication cohort for the benign vs any stage (stages I-IV) tumors. The solid black curve and shaded grey area as in (C), while the colored curves correspond to a subset of the samples split on histology. The label specifies the corresponding group and the point estimate of the AUC with the 95% confidence interval written out in paratheses. **(E)** As (D) but for a stratified analysis split on tumor stage. **(F)** Individual risk-scores from the model in (D) for the replication cohort. The vertical dashed line indicates the cut-off used.

**Table 3.** Proteins in multivariate models.

| Protein | Variable Importance<sup>a</sup> |  |  |
| --- | --- | --- | --- |
|  | Benign vs Stage I-II | Benign vs Stage III-IV | Benign vs Stage I-IV |
| WFDC2 | 0.819 | 0.939 | 0.905 |
| RBFOX3 | - | 0.918 | 0.872 |
| MUC16 | - | 0.911 | 0.867 |
| TCOF1 | - | 0.910 | 0.878 |
| KRT19 | 0.814 | 0.908 | 0.881 |
| L1RE1 | - | 0.892 | - |
| CRB2 | - | 0.864 | - |
| LEO1 | - | - | 0.843 |
| FOLR1 | 0.745 | - | 0.838 |
| BRK1 | 0.788 | - | - |
| VTCN1 | 0.746 | - | - |
| CRISP3 | - | - | 0.730 |

<sup>a</sup> A relative number on the scale of 0 to 1 ranking the individual importance of the proteins in each multivariate model. Rows in the table are sorted by the highest number across the three models.

**Table 4.** Predictive performance of the multivariate models.

| Model | Cohort | AUC | Sens<sup>a</sup> | Spec<sup>a</sup> |
| --- | --- | --- | --- | --- |
| Early | Disc | 0.85 | 0.96 | 0.24 |
|  | Repl | 0.93 | 1.00 | 0.16 |
|  | D vs R pval<sup>b</sup> | 0.21 | 1.00 | 0.25 |
| Late | Disc | 0.95 | 0.90 | 0.98 |
|  | Repl | 0.98 | 0.91 | 0.98 |
|  | D vs R pval<sup>b</sup> | 0.23 | 0.39 | 0.50 |
| Any | Disc | 0.96 | 0.95 | 0.67 |
|  | Repl | 0.96 | 0.97 | 0.68 |
|  | D vs R pval<sup>b</sup> | 0.79 | 0.52 | 0.43 |
|  | Repl (Early)<sup>c</sup> | 0.89 | 0.91 | 0.68 |
|  | Repl (Late)<sup>c</sup> | 0.97 | 0.98 | 0.68 |

<sup>a</sup> Sensitivity (Sens) and Specificity (Spec) achieved at a cut-off defined using the discovery cohort requiring at least 0.95 sensitivity. <sup>b</sup> P-values calculated with the DeLong's method for AUCs and Fisher's Exact test for Sens and Spec. <sup>c</sup> Point estimates of the performance of the 'Any stage' model when applied to Early- or Late-stage samples in the replication cohort.

## DISCUSSION

Detection of ovarian cancer is largely symptom driven, resulting in a considerable fraction of the cancer being discovered in late stages, which in turn leads to low 5-year survival rates. Far from all symptomatic women, however, are diagnosed with malignant tumors. Even at secondary centers in Sweden, when imaging techniques indicate adnexal ovarian mass, up to 75% of symptomatic women are surgically diagnosed with benign conditions*(8)*. The surgery itself is not risk-free and apart from complications related directly to the operations there are additional considerations regarding effects on fertility and induced menopause*(25)*. Further, if there is a strong indication of ovarian malignancy, the patient should be referred for surgery at a University hospital. Molecular tests that could complement, or replace, imaging techniques for separating benign from malignant conditions with high accuracy would benefit the patients themselves and provide an opportunity for health-care systems to reduce the workload for the highly trained experts, primarily at tertiary centers that interpret e.g. transvaginal ultrasounds. It has though, been difficult to find accurate enough biomarkers for ovarian cancer both with respect to screening and for triaging of symptomatic women.

In this study we employed high throughput ultra-sensitive affinity-based proteomics to perform unbiased searches for both univariate and multivariate biomarkers that could potentially separate malignant ovarian tumors from benign in symptomatic women. We found large differences in the plasma proteome between the benign and malignant groups and these effects were replicated in an independent cohort. We also concluded that only a small proportion of these protein biomarkers correlated with gene-expression in corresponding tumor tissue suggesting that the observable effects are possibly due to processes distinct from the developing tumor itself. This is in line with contemporary analyses of single-cell RNA expression*(5)* highlighting the role of the tumor microenvironment in relation to tumor progression*(6)*. In the sense of liquid biopsy-based biomarkers however, the ability to distinguish between diagnoses surpasses the need to know the exact origin of the protein. Indeed, here, only three out of the eight proteins in our multivariate signature displayed significant correlation with their respective mRNA expression in tumor tissue. In addition to replicating the performance of this multivariate signature in unseen samples with the AUC, we also validated the specific performance as indicated by sensitivity and specificity at a pre-determined cut-off level.

Our previously validated*(19)* multiplex protein biomarker signature was developed for the same clinical application as here, but with a much smaller number of available assays, 983 compared to the 5416 measured here. The previously developed signature combined 11 proteins out of which four (WFDC2, KRT19, MUC16, FOLR1) overlap with the 8-protein signature developed here. The remaining seven proteins were also measured with the assay used here, but the machine learning approach taken here prioritized other proteins over these. Comparing the performances of these two signatures is not completely straight forward, partly because of technical differences in the assay but primarily on how the cut-offs were developed. In terms of AUCs, our previous 11-panel achieved AUCs of 0.95 and 0.92 in the development and validation cohorts respectively, while our 8-protein panel here achieved AUCs of 0.96 in both cohorts. We previously used three cut-offs depending on intended use, one which focused on sensitivity, one for specificity and finally one providing the best balance between the two. Here, we focused on one type, requiring at least 95% sensitivity. At this cut-off, the performance in the replication cohort was estimated at a sensitivity of 97% with a specificity of 68%. Our 11-protein model was reported*(19)* to have a sensitivity of 97% at a specificity of 20% in the validation phase, calculated at the cut-off which was focused at sensitivity. It should be noted that this focused cut-off was developed for a target at 98% sensitivity so the comparison is not completely fair. In addition to an estimated higher performance in these measures, a major benefit is the lower number of proteins which facilitates a clinical implementation, both in terms of cost but also in terms of controlling variance and repeatability in the assay itself. In practice, several additional challenges apply when characterizing multiple plasma protein biomarkers. This includes, but are not limited to; a large expected dynamic range of the proteins in plasma which can be quite different from protein to protein, obtaining a high degree of multiplexing in the assays used while managing cost vs sample size and finally restrictions on the sample volume needed*(26)*. Despite these challenges, we have previously taken a multiplex assay developed with protein measured in NPX*(18)* to a validated*(19)* version with absolute quantification and these are necessary steps before applying the models here in clinical context. These steps also include retraining the models to fit the new concentration ranges and re-tuning of the cut-off to match the updated model.

The final model presented here for separating benign from malignant cases is based on eight proteins. Several of these have previously been associated with ovarian cancer, either on expression as characterized at gene or protein level (WFDC2, MUCIN16, KRT19*(19)*, RBFOX3*(22)*, CRISP3*(27)*) or as drug targets (FOLR1*(28)*), while for instance TCOF1*(29)* have been implicated in other cancers but, to the best of our knowledge, not with ovarian cancer. The last protein in the model, LEO1, has been shown*(30)* to regulate heterochromatin with downstream importance in maintenance of cellular quiescence. Overall, our results indicate that it would be difficult to carry out a specific pre-selection of what potential biomarkers to characterize either based on previous indications on protein level or based on gene-expression in tumor tissue. Here, only 21% of the potential single valued protein biomarkers showed a strong correlation between tumor gene expression and plasma protein concentrations while the vast majority showed no correlation. One such example is the RBFOX3 protein, which was one of the highest ranked single biomarkers for separating benign from malignant tumors as well as ranked with very high importance in the multivariate models, albeit with weak correlation with its corresponding gene expression (R = 0.11). When examining the protein-protein correlation networks, we found RBFOX3 to be part of a larger network of highly correlated plasma proteins out of which 6 other members did show correlation with their corresponding gene expression.

This suggests that a large proportion of the potential plasma biomarkers presented here could be downstream effects of the growing tumor or from the tumor microenvironment rather than an underlying driver of the tumor development. This is highly relevant and put restrictions on the interpretation of the proteins in relation to cancer biology and also restricts the possible use of these protein biomarkers as future drug-targets, at least until their role in relation to the disease is understood.

Our study has several limitations. We are limited by the number of samples analyzed in the sense that specific stratification of separate histology in combination with e.g. stage or other clinical parameters are not possible given the current material. Although geographically separate, both our cohorts are Swedish and further studies in groups of other ethnicities would have been beneficial to better understand the variation. Neither have we evaluated the models in healthy women, such analyses could give insights into any future application in screening and to evaluate variation in the scores in relation to anthropometrics, life-style variables, or genetics, which have all been shown to affect circulating plasma protein levels*(31–33)*.

In conclusion, our study encompasses the largest plasma proteome study to date in relation to ovarian cancer. Our results include a large number of replicated plasma protein biomarkers for separation of benign and malignant tumors in symptomatic women where we also show that only a fraction of these correlate with tumor gene expression. A multivariate model containing eight proteins showed excellent replicated performance in separating benign from malignant tumors regardless of tumor histology and thus could have clinical use as a triaging tool for symptomatic women.

## MATERIALS AND METHODS

### Study design

This study included clinical samples collected from two geographically separated locations in Sweden. Both cohorts consisted of women that have been surgically diagnosed with either benign or malignant tumors after suspicion of ovarian cancer. Samples were collected according to standardized protocols and according to local regulations. All participants gave written consent, and all necessary ethical permits were in place. One cohort was used strictly as discovery and the second as validation cohort. A high-throughput proteome assay was used to characterize the plasma proteome in both cohorts at the same time in the same laboratory. RNA-sequencing was used to characterize gene expression in fresh frozen tumor tissue from a subset of the women in the validation cohort. Protein biomarkers were analyzed both individually and in combination in relation to clinical endpoints. Strict adjustment for multiple hypothesis testing was used throughout.

### Samples

Plasma samples of women with benign and malignant ovarian tumors were collected from the tumor biobank*(34)* at Biobankvast.se, Western healthcare region, Göteborg, Sweden (N=171) and from the U-CAN collection*(35)* at Uppsala Biobank, Uppsala University, Sweden (N=233). Inclusion criteria was suspicion of ovarian cancer followed by surgical diagnosis of either malignant or benign conditions. Exclusion criteria were patients that had received neoadjuvant treatment prior to surgery or if the tumor was pathologically determined to be metastases originating from other tissues. The samples from Göteborg were collected from 2016 to 2018 and the samples from U-CAN in Uppsala between 2012 and 2018. All samples were collected in agreement with local guidelines and regulations. The tumors were examined by pathologist specialized in gynecologic cancers for histology, grade, and stage according to International Federation of Gynecology and Obstetrics (FIGO) standards. Both cohorts contained mixed tumor histology. In the Göteborg samples, 70.6% were high grade serous (HGSC), 8.2% low grade serous (LGSC), 7.0% mucinous, and the remainder clear cell, endometroid, sarcoma, epithelial/clear cell, mucinous/teratoma or unclear histology. In the U-CAN samples, among the samples with complete histology data, 60.1% were HGSC, 8.7% LGSC, 7.6% endometroid, 6.0% clear cell, 5.5% carcinosarcoma and the remainder mucinous, non-epithelial, endometroid or mixed. All plasma samples were frozen and stored at −70°C. Basic statistics for the samples used are presented in Table 1. The study was approved by the Regional Ethics Committee in Uppsala (Dnr: 2016/145) and Göteborg (Dnr: 201-15) and informed written consent was obtained from all participants following the guidelines of the Declaration of Helsinki.

### Plasma protein characterization

The plasma proteome was analyzed using the proximity extension assay (PEA) as implemented in the Explore HT-version (Olink Proteomics AB, Uppsala, Sweden). The samples were analyzed at the Olink Service Laboratory in Uppsala, Sweden. In brief, the PEA is based on pairs of antibodies equipped with probes, DNA single-strand oligonucleotide reporter molecules, that bind to their respective target if present. Target binding by both probes in close proximity generates double-stranded DNA amplicons which are then quantified by next generation sequencing*(36)*. Here, 5616 unique proteins were characterized in each of the 404 samples. The samples were randomized across plates with respect to cohort and diagnose (benign or malignant). In the resulting data-file provided by the analysis platform, each individual protein measurement, assay and sample is labelled depending on passing or failing quality control as provided by the instrument software. Here, a total on 768 individual measurements (corresponding to 0.036%, 768/(404*5416)) did not pass quality control and were removed from further analyses. Two proteins (Apolipoprotein E (APOE) and Fibronektin 1 (FN1)) had detection rates below 95% across all samples and were removed from further analyses. The resulting NPX values are on a log2 scale and in the logarithmic phase of the curve, one (1) increase of the NPX value corresponds to a doubling of the protein content. In the resulting data, a high NPX value corresponds to a high protein concentration. Each of the measured proteins has a lower limit of detection (LOD) given in the same NPX-scale which is determined at run time. Here, measurements under LOD were kept as is in the downstream analysis. After quality control, the detection rate across all samples were 99.5 to 99.8%.

### Tumor RNA extraction

Fresh frozen tumor tissue samples from women in the Uppsala cohort was used for analysis of mRNA expression. Nucleic acids were isolated using the AllPrep® DNA/RNA Micro Kit (Cat.no. 80284, Qiagen, Hilden, Germany). From the fresh frozen tumor samples, approximately 2-5 cryosections (thickness 10 µm) were obtained, using a CryoStar NX70 Cryostat (Thermo Fisher Scientific™ Inc.). Cryosections were immediately transferred into 1,5 ml Eppendorf tubes containing 600 μl Buffer RLT with addition of 1% β-Mercaptoethanol (Cat.no. M6250-10ML, Merck, Darmstadt, Germany). Samples were homogenized by continuous vortexing for 30 seconds, followed by simultaneous extraction of total RNA and genomic DNA from each sample, according to the manufacturers protocol for microdissected cryosections. All RNA samples were subjected to DNase treatment using the RNase-Free DNase Set (Cat.no. 79254, Qiagen, Hilden, Germany), and finally eluted in RNase free water before storage at -80°C. RNA yield and integrity were measured on the Agilent 2100 system, using the RNA 6000 Nano Kit (Cat.no. 5067-1511, Agilent Technologies, Santa Clara, CA, USA).

### mRNA sequencing and alignment

Sequencing libraries were prepared from 122-194 ng (three samples), 200 ng (seven samples) or 500 ng (71 samples) total RNA using the TruSeq stranded mRNA library preparation kit (cat# 20020595, Illumina Inc.) including polyA-selection. Unique dual indexes (cat# 20022371, Illumina Inc.) were used. The library preparation was performed according to the manufacturers’ protocol (#1000000040498, Illumina Inc.). The libraries were then sequenced on a NovaSeq 6000 system (Illumina Inc.) on S4 flowcells with version 1.5 sequencing chemistry on three lanes. Paired-end sequencing of with read lengths of 150 bp was used. Across all sequenced samples,112 to 477M raw reads were obtained. The data was analyzed with the nf-core framework*(37)* version 3.3 (https://nf-co.re/rnaseq/3.3). In brief, the reads were aligned to human reference genome (GRCh38) using the STAR software suite and 82 to 221M aligned reads were obtained for each sample. The Ensembl database was used for annotation of genes and the TPM (transcript per million) value was used as representation of gene expression.

### Statistics and data analysis

All analyses were carried out in R (version 4.2.3) *(38)*. The univariate comparisons were done one protein at a time using a two-sided Wilcoxon ranked based test. The resulting p-values were adjusted for multiple hypothesis testing using the Holm correction method as implemented in the ‘p.adjust’ R-function. The correlations between each plasma protein and corresponding tumor gene expression were calculated using the ‘cor.test’ function in R with the Spearman method and resulting p-values adjusted using the Holm method as above. The protein-protein correlations were calculated using the Spearman method via the ‘cor.test’ function in R. This analysis was restricted to protein pairs in which at least one protein was among the 191 univariate significant proteins, and the p-values were adjusted using Holm’s method as above. Using proteins that had significant (q < 0.05) absolute correlations of at least 0.8 in the discovery cohort, the network and clusters were built using the ‘igraph’ R package*(39)* with clusters identified using the Leiden method based on modularity. In the visualization, network nodes were scaled between 1 and 20, and the clusters between 3 and 50, by the number of degrees. Prior to the multivariate analysis, NPX-values were normalized between cohorts by first calculating a per-protein normalization factor as the difference of the mean of the benign between the discovery and the replication cohort. This normalization factor was then added to each individual measurement in the replication cohort. Multivariate models were built using the discovery cohort only. Three separate models were created: early-stage (I and II), late-stage (III and IV) and any stage ovarian cancer (I-IV) versus benign, respectively, in the same way. Starting from the 191 univariately significant proteins, a feature selection step was first done by recursive feature elimination as implemented in the ‘rfe’ function in the ‘caret’ R-package*(40)*. Distance weighted discrimination*(24)* with a polynomial kernel from the ‘kerndwd’ (version 2.0.3) R package*(41)* was then used to create a prediction model. A tuning step was performed during training over the ‘lambda’, ‘qval’, ‘degree’ and ‘scale’ parameter. The final three models used the following sets for these parameters early stage: (0.01, 0.05, 1, 0.1), late stage: (0.1, 0.05, 1, 0.1) and any stage: (0.1, 0.05, 3, 0.1). The output from the model was risk-score on the scale 0 to 1 and ROC-curves was generated using the ‘pROC’ R-package*(42)*. A cut-off for malignancy was then developed using the ROC-curve from the discovery cohort and taken at the first point on the curve with at least 95% sensitivity. The model was then applied to the replication samples and a decision was made based on the cut-off from the discovery cohort. Obtained areas under the curve (AUCs) were compared between the discovery and replication cohort using the ‘roc.test’ function from the ‘pROC’ R-package*(42)* with the DeLong’s method. Obtained sensitivities and specificities at the cut-off was compared between the discovery and replication cohorts using a Fisher’s exact test.

## List of Supplementary Materials

Tables S1 to S4 are given in the supplementary material.

## Data Availability

All data produced in the present study are available upon reasonable request to the authors.

## Acknowledgments

Sequencing was performed by the SNP&SEQ Technology Platform in Uppsala. The facility is part of the National Genomics Infrastructure (NGI) Sweden and Science for Life Laboratory. The SNP&SEQ Platform is also supported by the Swedish Research Council and the Knut and Alice Wallenberg Foundation. The computations and data handling were enabled by resources in project sens2023504 provided by the National Academic Infrastructure for Supercomputing in Sweden (NAISS) at Uppsala Multidisciplinary Center for Advanced Computational Science (UPPMAX) partially funded by the Swedish Research Council through grant agreement no. 2022-06725. Additional support was provided by Paul-Theodor Pyl from the Swedish Bioinformatics Advisory Program at the National Bioinformatics Infrastructure Sweden (NBIS).

## Funding

The Swedish Research Council 2022-00857 (SE)

The Swedish Cancer Foundation 220604FE (SE)

The Swedish Cancer Foundation 232874PJ (SE)

The Swedish Cancer Foundation 190008PJ (UG)

The Swedish Cancer Foundation CAN211848 (KSu)

The Swedish state under the agreement between the Swedish government and the county council, the ALF-agreement (KSu)

## Author contributions

Conceptualization: UG, KSu, KSt, SE

Data curation: EI, SP, SE

Formal analysis: MM, SE

Funding acquisition: UG, KSt, KSu, SE

Investigation: MM, JHL, SP, ML, EI, AT, UG, KSu, KSt, SE

Methodology: MM, SE

Project administration: JHL, SE

Resources: AT, KSu, KSt

Software: MM, SE

Supervision: UG, SE

Visualization: MM, SE

Writing – original draft: MM, SE

Writing – review & editing: All authors

## Data and materials availability

Raw data for the RNA-sequencing is being deposited in the Federeated Europeean Genome Archive (FEGA) but has not yet received an access number. The proteomics data will be made available in the Swedish Science for Life Laboratory Data Repository and through that resource available to researchers on reasonable request.

